# A Comparative Analysis of System Features Used in the TREC-COVID Information Retrieval Challenge

**DOI:** 10.1101/2020.10.15.20213645

**Authors:** Jimmy Chen, William R. Hersh

## Abstract

The COVID-19 pandemic has resulted in a rapidly growing quantity of scientific publications from journal articles, preprints, and other sources. The TREC-COVID Challenge was created to evaluate information retrieval methods and systems for this quickly expanding corpus. Based on the COVID-19 Open Research Dataset (CORD-19), several dozen research teams participated in over 5 rounds of the TREC-COVID Challenge. While previous work has compared IR techniques used on other test collections, there are no studies that have analyzed the methods used by participants in the TREC-COVID Challenge. We manually reviewed team run reports from Rounds 2 and 5, extracted features from the documented methodologies, and used a univariate and multivariate regression-based analysis to identify features associated with higher retrieval performance. We observed that fine-tuning datasets with relevance judgments, MS-MARCO, and CORD-19 document vectors was associated with improved performance in Round 2 but not in Round 5. Though the relatively decreased heterogeneity of runs in Round 5 may explain the lack of significance in that round, fine-tuning has been found to improve search performance in previous challenge evaluations by improving a system’s ability to map relevant queries and phrases to documents. Furthermore, term expansion was associated with improvement in system performance, and the use of the narrative field in the TREC-COVID topics was associated with decreased system performance in both rounds. These findings emphasize the need for clear queries in search. While our study has some limitations in its generalizability and scope of techniques analyzed, we identified some IR techniques that may be useful in building search systems for COVID-19 using the TREC-COVID test collections.

## INTRODUCTION

Since the World Health Organization declared the Coronavirus Disease 2019 (COVID-19) a public health emergency,^1^ there has been explosive growth in scientific knowledge about this novel virus. Consequently, the use of preprints and fast-track publication policies has resulted in a significant increase in the number of COVID-19 related publications over a short period of time.^2,3^ Information retrieval (IR, also known as search) systems are the tool usually employed to manage access to large corpora of literature.^4^ The efficacy of IR systems is often assessed in challenge evaluations that provide reusable test collections, such as those led by the National Institutes of Standards and Technology (NIST) in the Text REtrieval Conference (TREC).^5^

To address the need for system evaluation in this rapidly changing information environment, NIST sponsored the TREC-COVID Challenge.^6^ As with most IR challenge evaluations, test collections of documents, topics for searching, and relevance judgments were developed.^7^ The document collections were derived from snapshots of the COVID-19 Open Research Dataset (CORD-19), a regularly updated dataset of manuscripts consisting of coronavirus-related research gathered from various sources including journal articles, PubMed references, arXiv, medRxiv, and bioRxiv.^3^ Based on the the TREC framework, the TREC-COVID Challenge tasked researchers to evaluate IR systems over 5 rounds to retrieve manuscripts relevant to topics about COVID-19 from CORD-19, with the goal of building a reusable test collection for further research.^8^ Every 2-3 weeks, researchers implemented and evaluated retrieval systems with un updated CORD-19 dataset and the addition of new search topics.^9^ While there exists previous work examining techniques used in other IR test collections,^10–12^ there have not yet been any studies comparing the methods and systems used by participants in the TREC-COVID Challenge.

The purpose of this study was to compare performance in different approaches used in the TREC-COVID Challenge by: 1) developing a taxonomy to characterize IR techniques and system characteristics used, and 2) applying this taxonomy to identify features of IR systems associated with higher performance. Using run reports from Round 2 and Round 5, we designed a taxonomy and evaluated its features using a univariate and multivariate regression analysis. In this study, we assessed how certain methodologies were associated with higher retrieval performance and discussed the implications and limitations of our analysis.

## METHODS

### Dataset and Features

The TREC-COVID challenge^6^ occurred over 5 rounds in 2020 on the rapidly growing CORD19 dataset,^3(p19)^ with 30 initial topics in Round 1 and 5 new topics each subsequent round. Each topic consisted of three fields: (1) a short *query* statement that a user might enter, (2) a longer *question* field more thoroughly expressing the information need of the topic, and (3) a *narrative* field describing what would constitute a relevant document. After Round 1, relevance judgments, consisting of IDs of manuscripts assessed by human assessors as relevant, partially relevant, or irrelevant, were made available for previously unassessed manuscripts after each round. **Table 1** summarizes the CORD19 corpora, topics, number of teams, runs, and judgments present in each round of the TREC-COVID challenge.

**Table 1.** Overview of the TREC-COVID challenge. Over 5 rounds, research teams implemented information retrieval (IR) systems to search the growing CORD-19 dataset. After each round, new topics and relevance judgments of manuscripts from previous iterations of the CORD-19 dataset were released for use in subsequent rounds.

| Round | CORD-19<br>Date | Documents | Docs<br>Changed | Topics -<br>new/total | Teams | Runs | Cumulative<br>Judgments |
| --- | --- | --- | --- | --- | --- | --- | --- |
| 1 | 4/10/2020 | 51103 | N/A | 30/30 | 56 | 143 | 8691 |
| 2 | 5/1/2020 | 59851 | 20 | 5/35 | 51 | 136 | 20728 |
| 3 | 5/19/2020 | 128492 | 2017 | 5/40 | 31 | 79 | 33068 |
| 4 | 6/19/2020 | 157817 | 104 | 5/45 | 27 | 72 | 46203 |
| 5 | 7/16/2020 | 191175 | 1137 | 5/50 | 28 | 126 | 69318 |

Reports for all submitted runs, including those from Rounds 2 and 5, were made publicly available from the TREC-COVID Challenge (https://ir.nist.gov/covidSubmit/archive.html) and were manually reviewed by J.C. We chose to review reports from Rounds 2 and 5 because we wanted to compare methodologies used in two different rounds where feedback methods from topics from previous rounds were available. Each run report was written as a textual description of the methodology used to produce the run in whatever detail the submitting team provided. An example run report is shown **(Figure 1)**.

**Figure 1.**
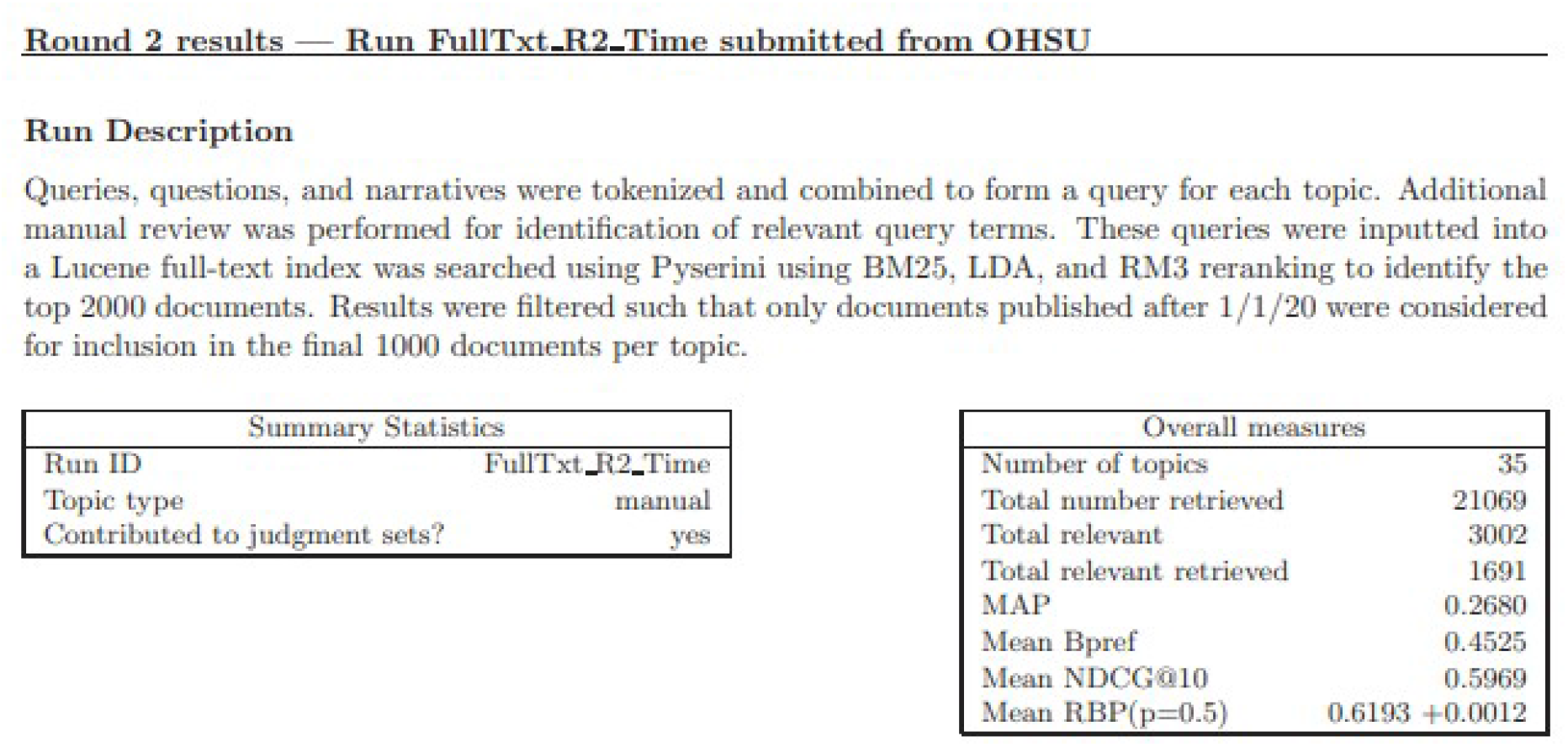
Example Run Report from Round 2. During submission of a run, participants were encouraged to provide a methodological description of each submitted run. This run description, along with the run ID, topic types, and performance metrics, were reported in a publicly available repository of archived results (https://ir.nist.gov/covidSubmit/archive.html). These run reports were manually reviewed for features.

The following features were extracted for each run in the reports from Round 2 and Round 5:

- Text used (i.e., title and abstract only, paragraph-based indices, or full-text)
- Type of query (i.e., any combination of the query, question, and narrative from the TREC-COVID topic fields)
- Any query pre-processing (i.e., stemming, removing stop words)
- Query term expansion (addition of terms not originally provided in each topic)
- Manual review methods (i.e., human interventions including the use of human assessors in Continuous Active Learning)^13^
- Any weighted ranking system used (i.e. non-neural scoring functions such as BM25^14^ and term frequency–inverse document frequency, or TF-IDF^15^)
- Any ranking model that used a neural architecture (including deep transformer models such as BERT,^16^ SciBERT,^17^ T5^18^ as well as DeepRank,^19^ a neural network that attempts to simulate humans in relevance judgments)
- Other techniques (machine learning models such as SVM, logistic regression, custom scoring functions, otherwise known as term proximity scores. Custom search methods were also included in this category including ReQ-ReC,^20^ a double-loop retrieval system)
- Dataset used to fine tune any system (i.e., MS-MARCO, a large dataset of annotated documents based off 100,000 Bing queries,^21^ CORD-19 dataset transformed into document vectors, and relevance judgments from previous rounds)
- Fusion of multiple runs into a single run (including use of reciprocal rank fusion,^22^ COMB fusion methods^23^)
- Re-ranking implemented, defined as whether a second system (most commonly a neural network) was used to refine an initial scoring system.
- Pseudo-relevance feedback, or system-generated relevance feedback based on an initial query.
- How/if human-generated relevance feedback, or relevance judgements, from the previous round(s) were used.
- Runs filtered by date. Removing documents published before 2020 (or when the pandemic began to gain widespread notice) had been previously suggested by McAvaney et al. in their post-hoc analysis of their neural re-ranking system as a possible method to improve performance.^24^

These features were selected to encompass a broad set of commonly used techniques in ad-hoc retrieval. Of note, TREC challenges typically do not occur over multiple rounds; thus, the addition of relevance judgments was a novel addition to the TREC-COVID challenge. Since the length of reports varied at the researcher’s discretion, many reports likely had some number of missing features. To minimize the impact of these null features, we assumed that runs that did not provide information about the type of text or query used in their system likely searched on the full-text using the query subfield from each topic. Features extracted from reports were either characterized as a binary feature (used or not used in the system) or a categorical feature (a description of feature for the system, i.e., BM25 as the weighted system used). Categorical features were later one-hot encoded, or converted into binary features over multiple columns, prior to input into our regression analysis. The extracted features and their encoding is shown in **Table 2**

**Table 2.** Taxonomy Features Extracted from Run Reports. Features extracted from run reports from Round 2 and Round 5 are listed. Features were either extracted as a binary feature or a categorical feature (in which a short description specifying the feature was provided). Prior to input into univariate and multivariate analysis, categorical features were one-hot encoded, or converted into binary variables over multiple columns.

| Feature Names | Input Type | No. of Binary Values > 0 |  |
| --- | --- | --- | --- |
|  |  | Round 2 | Round 5 |
| Title + Abstract Index Used | Binary | 42 | 78 |
| Paragraph Index Used | Binary | 29 | 27 |
| Full-Text Index Used | Binary | 92 | 70 |
| Filtered Dataset by Time | Binary | 8 | 5 |
| Query | Binary | 109 | 111 |
| Question | Binary | 56 | 60 |
| Narrative | Binary | 28 | 32 |
| Query Preprocessing | Binary | 29 | 42 |
| Term Expansion | Binary | 37 | 23 |
| Manual Review | Binary | 21 |  |
| Weighted System | Categorical<sup>a</sup> | N/A | N/A |
| Neural | Binary | 49 | 67 |
| Dataset for Fine-Tuning | Categorical<sup>a</sup> | N/A | N/A |
| Other Technique | Categorical<sup>a</sup> | N/A | N/A |
| Re-ranking | Binary | 44 | 68 |
| Fusion Technique Used | Binary | 31 | 111 |
| Pseudo-relevance feedback | Binary | 44 | 24 |
| Feedback from Judged Manuscripts | Binary | 23 | 59 |

<sup>a</sup>Categorical features were features that were extracted as a description rather than a binary variable.

We included all runs that contained more than 1 extracted feature to ensure a reasonably large enough and useful dataset for analysis. We excluded unusually poorly performing runs that likely represented poor system or method implementations. The exclusion threshold for these runs was defined as an average performance of less than 0.2 across all 5 performance metrics used to evaluate run performance in the TREC-COVID Challenge. Performance metrics reported by NIST and used in our analysis included: precision at K documents (P@K), normalized discounted cumulative gain at K documents (NDCG@K), rank-based precision with depth = 5 (RBP (p = 0.5)), binary preference (bpref), and mean average precision (MAP). In Round 2, the depth of documents for P@K and NDCG@K were 5 and 10 respectively, while in Round 5, the depth of documents for P@K and NDCG@K were both 20. These changes were made out of concern for inflated performance when evaluating precision on a small number of documents.^6^ For each run, these performance metrics were computed as the mean performance across all topics in the round.

### Univariate and Multivariate Analysis

All data analysis and pre-processing was performed using R (version 4.0.2)^25^ using the glmnet package.^26^ For each round, 5 univariate linear regressions were created using all extracted features as the independent variables, and each of the 5 performance metrics (NDCG@K, P@K, RBP, bpref, and MAP) as the dependent variable. Coefficients and standard errors were calculated for each feature, and p-values were extracted for each feature coefficient, with significance defined as p < 0.05. Features that met the threshold for significance in the univariate regression were subsequently input into a multivariate linear regression. Overall, positive coefficients were interpreted to be associated with higher performance. Therefore, features which remained significant after both univariate and multivariate regression were likely associated with high performance in the TREC-COVID challenge.

## RESULTS

Round 2 consisted of 136 runs submitted by 51 teams (with a permitted maximum of 3 runs submitted per team), and Round 5 consisted of 126 runs submitted by 28 teams (with a permitted maximum of 8 runs submitted per team). The topics in Round 2 included 30 previous (i.e., from Round 1) topics with relevance judgments, and 5 new topics. The topics in Round 5 included 45 previous (i.e., from Rounds 1-4) topics and 5 new topics. Overall, 110 runs from 42 teams met inclusion criteria in Round 2 and 111 runs from 23 teams met inclusion criteria in Round 5. The proportion of manual (defined as involving human intervention), feedback (defined as using judgments from prior rounds), and automatic (defined as neither feedback nor manual) runs varied between runs. In Round 2, the majority of the runs were categorized as automatic runs; in round 5, the majority of the runs were characterized as feedback runs. These findings are summarized in **Table 3**

**Table 3.**
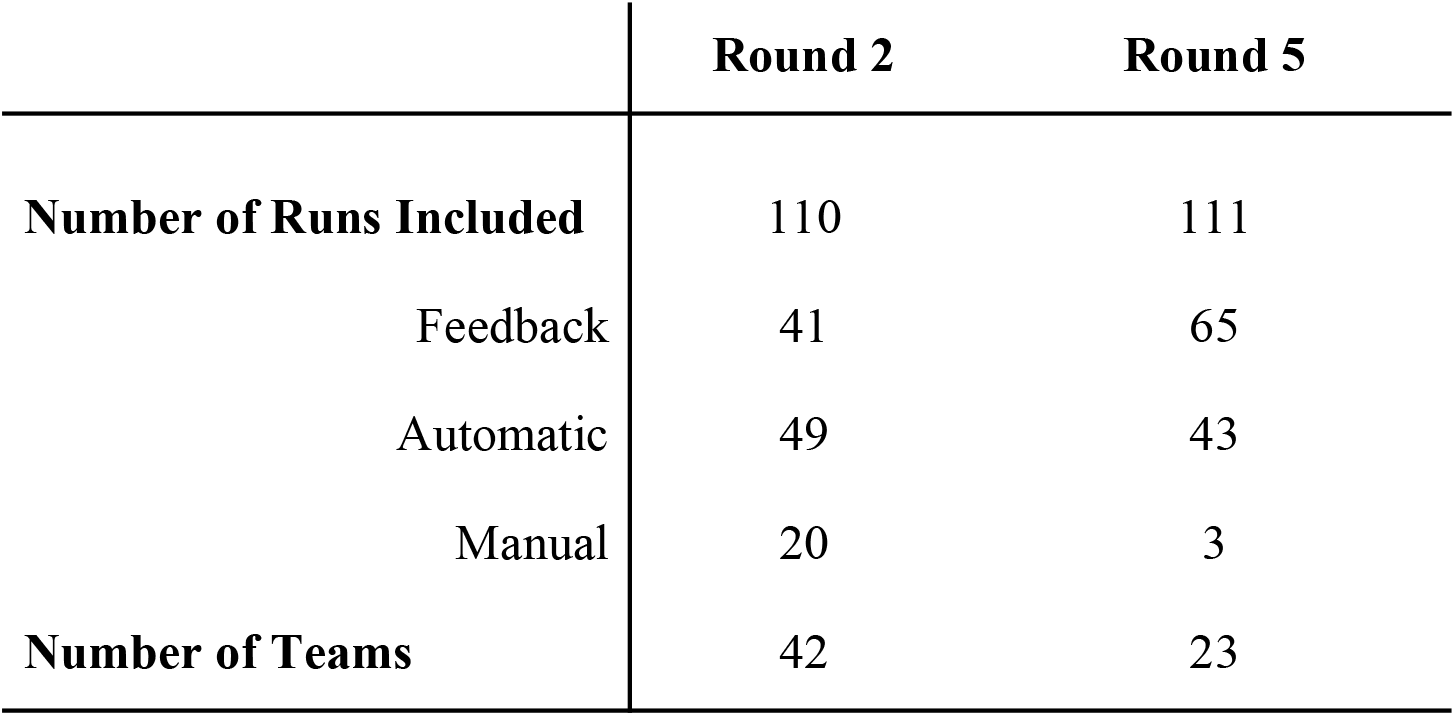
Distribution of Included Runs in Rounds 2 and 5. The number of included runs and the number of participating teams is included in this table. Runs were either sub-categorized as feedback (defined as using judgments from prior rounds), manual (involving human intervention), and automatic (neither manual nor feedback) runs.

|  | Round 2 | Round 5 |
| --- | --- | --- |
| **Number of Runs Included** | 110 | 111 |
| Feedback | 41 | 65 |
| Automatic | 49 | 43 |
| Manual | 20 | 3 |
| **Number of Teams** | 42 | 23 |

Significant features for the 5 univariate regressions each for Round 2 and Round 5 are shown in **Figure 2** and varied depending on the performance metric used. In Round 2, query term expansion (n = 37 runs), fine-tuning of ranking systems on MS-MARCO (n = 18 runs), Round 1 judgments (n = 9 runs), or document vectors formed by the CORD-19 dataset (n = 9 runs) were associated with higher performance across most, if not all, performance metrics. Use of ReQ-Rec (n = 3 runs submitted by 1 team), and narrative text in the query (n = 28 runs) were associated with decreased performance across the majority of performance metrics. In Round 5, use of the question text in the query (n = 32 runs) and TF-IDF vectors were associated with increased performance (n = 14 runs), whereas the use of neural networks, narrative text in the query (n = 67 runs), and proximity score (n = 2 runs) were associated with decreased performance across all performance metrics.

**Figure 2.**
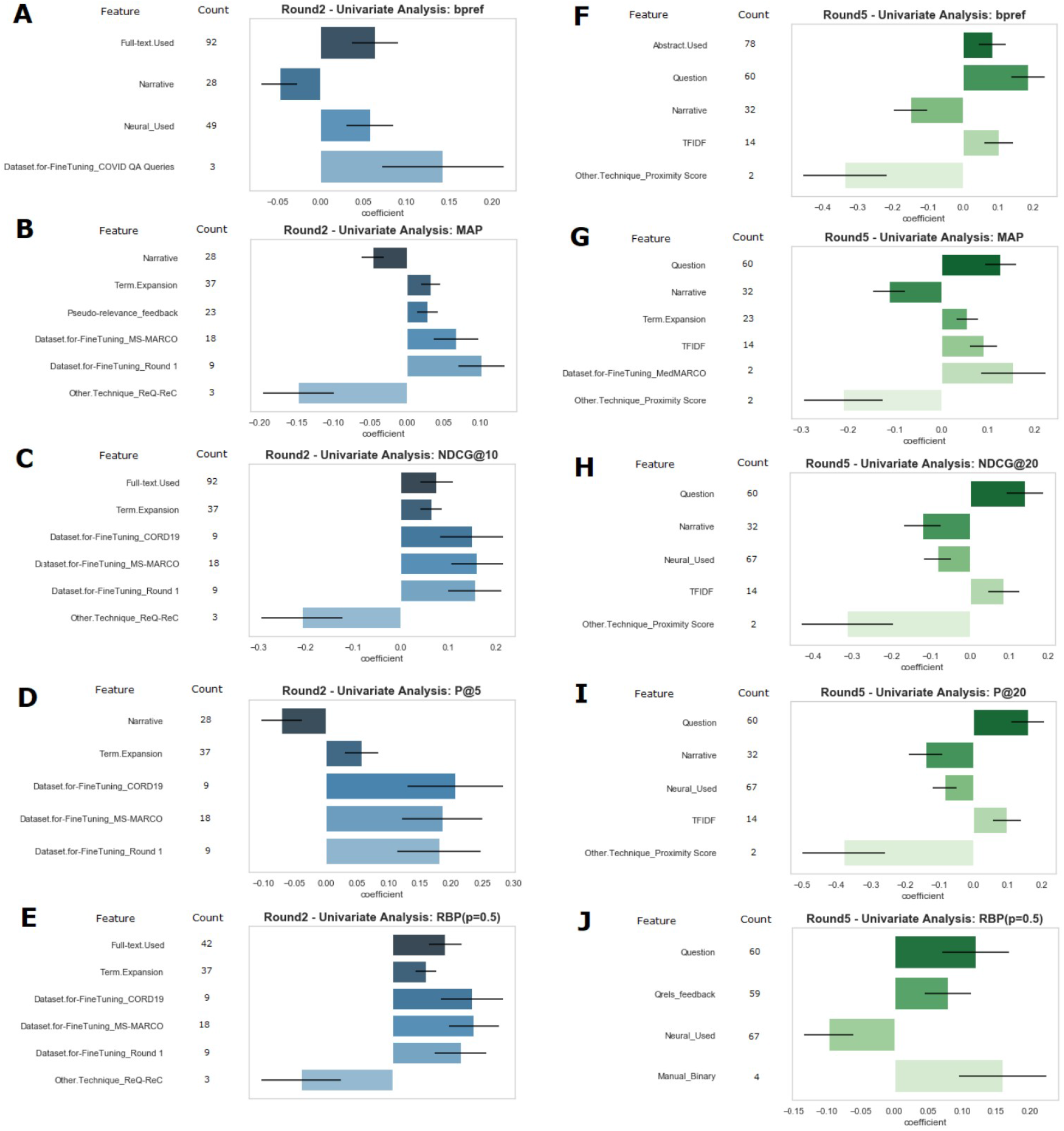
Significant Features after Univariate Regression Analysis in Rounds 2 and 5. Univariate analysis was performed on features extracted from reports from Rounds 2 and 5. Features that were significant after input into a univariate linear regression are shown for the following performance metrics from Rounds 2 and 5 respectively: binary preference (**A** and **F**), mean average precision (**B** and **G**), normalized distributive cumulative gain (**C** and **H**), precision @ k documents (**D** and **I**), and rank-based precision (**E** and **J**). The count, or number of times that the feature occurred in our extracted dataset, is displayed adjacent to the feature name. These significant features were subsequently input into a multivariate regression to determine which features were independently associated with performance.

Significant features from multivariate regressions on the 5 different performance metrics in Rounds 2 and 5 are shown in **Figure 3**. After features found to be significant on univariate regression were input into a multivariate regression, the following features remained significantly associated with increased performance in Round 2 with the majority of performance metrics: term expansion (n = 37), ranking system fine-tuning on CORD-19 vectors (n = 9), MS-MARCO (n = 18), and Round 1 judgments (n =9). Using ReQ-Rec (n = 3) remained significantly associated with decreased performance. In Round 5, using the question text to formulate the query (n = 60) and TF-IDF vector weighting (n =14) were associated with increased performance, while a custom proximity score (n = 2) as a scoring function was associated with decreased performance. As seen in Round 2, using feedback in Round 5 (n = 59) was associated with increased performance when runs were evaluated on RBP.

**Figure 3.**
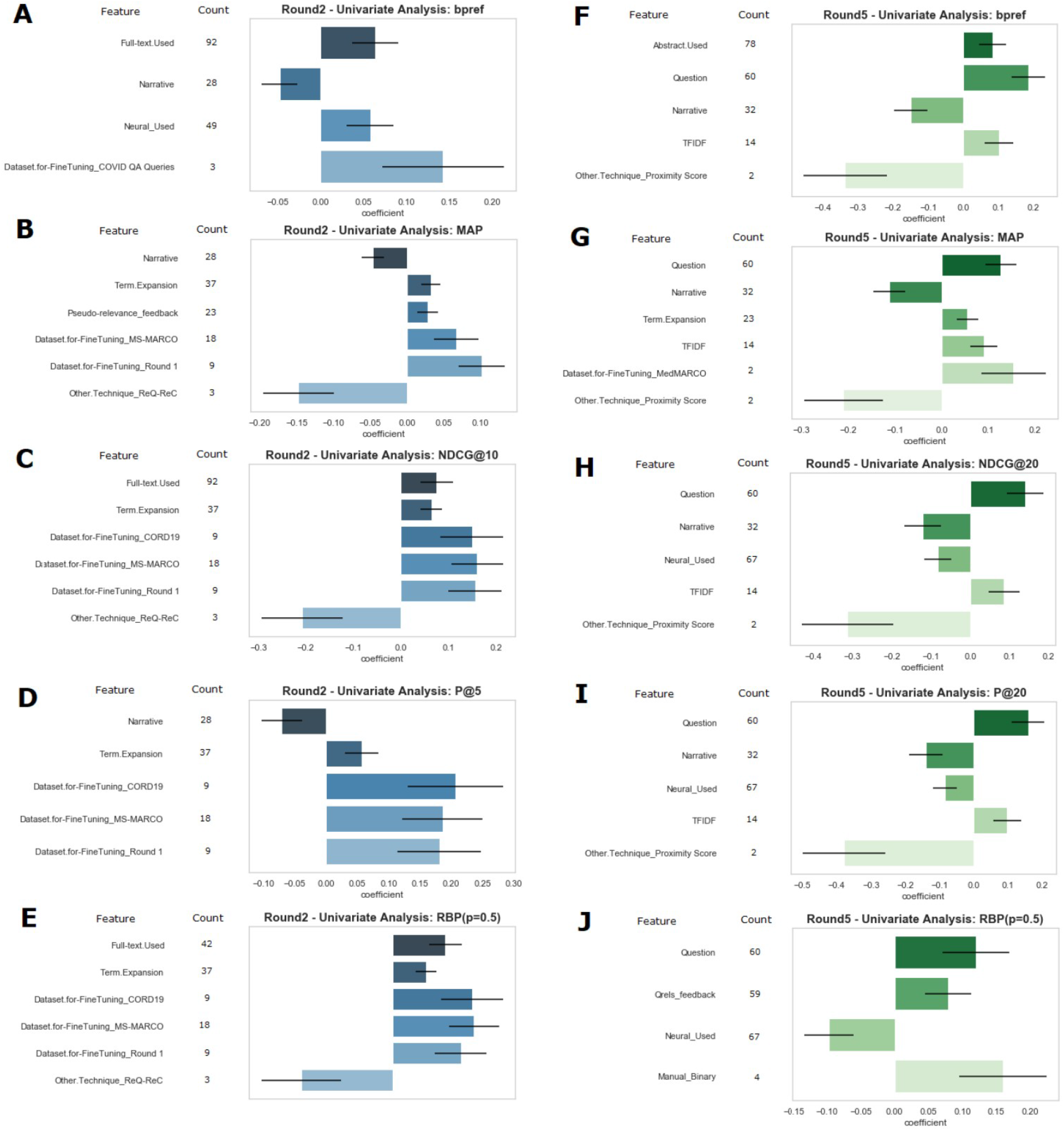
Significant Features after Multivariate Regression Analysis in Rounds 2 and 5. Features that were found to be significant in univariate regression were further input into a second, multivariate regression. Significant features were reported for the following performance metrics in Rounds 2 and 5 respectively: binary preference (**A** and **F**), mean average precision (**B** and **G**), normalized distributive cumulative gain (**C** and **H**), precision @ k documents (**D** and **I**), and rank-based precision (**E** and **J**). Depending on the coefficients, these features were concluded to be significantly associated with increased or decreased performance in the TREC-COVID challenge.

## DISCUSSION

This study aimed to develop a taxonomy of features to evaluate techniques associated with higher performance in runs submitted to Rounds 2 and 5 of the TREC-COVID Challenge. The key findings were: 1) fine-tuning ranking systems using relevance judgments resulted in significant improvement in performance, particularly in Round 2, and 2) query formulation is an important component of successful search.

Our first key finding was that fine-tuning ranking systems using relevance judgments resulted in significant improvement in performance, particularly in Round 2. Unlike previous TREC challenges, rapid turnout of relevance judgments over multiple rounds resulted in opportunities to fine-tune ranking systems for improved performance. Many of the runs labelled as feedback runs (n = 41 in Round 2 and n = 65 in round 5) employed fine-tuning, though a small portion specifically used the relevance judgments specifically in fine-tuning their ranking systems. Other teams who fine-tuned on similar datasets, including the vectorized CORD19 dataset (represented as Dataset.for-FineTuning_CORD19) and MS-MARCO also achieved comparable levels of improvement when compared to systems that did not use fine-tuning on these specific datasets. The noted improvement of fine-tuning on an annotated dataset has been reported in other TREC challenges, most notably the usage of MS-MARCO by Nogueira et al to refine a neural network system that vastly outperformed other runs in the TREC CAR challenge.^27^ Interestingly, the benefits of fine-tuning systems did not persist into Round 5 (with the exception of evaluation on RBP) despite more prevalent use of neural systems and feedback runs.

Since the TREC-COVID Challenge brought together a mix of research teams with varying experience in IR challenge evaluations, along with the short time between rounds (2-3 weeks), the absence of significance with fine-tuning on previous round judgments may be explained by implementation differences between teams, as many teams implemented variations of the popular sequence of an initial weighted system (most commonly BM25) followed by a neural re-ranker (i.e., BERT with or without fine-tuning on MS-MARCO or previous relevance judgments).^28,29^ However, since we one-hot encoded other techniques, our linear regressions may have overrepresented individual techniques that few teams used (including ReQ-ReC in Round 2, and proximity scoring in Round 5). Future work may be needed to validate the performance of other techniques compared with the standard weighted and neural pipelines. Furthermore, since we chose to set neural networks as a binary variable, there may be opportunities to explore how different architectures influence performance in the TREC-COVID challenge.

Our second key finding was that query formulation was an important component of successful search. While most teams used the query and question fields in formulating an input query, several teams (n = 28 and 32 in Rounds 2 and 5 respectively) chose to use the narrative portion of the topic, which was associated with decreased performance in both rounds. Because the narrative contained freehand descriptions qualifying each topic, these descriptive fields were noisy. For example, topics 33 and 34 contained the phrase “excluding…,” with subsequent wording describing what not to search. Furthermore, vocabulary used in the topics designed by the TREC-COVID organizers were not consistently used in manuscripts included in the CORD-19 dataset (i.e. differences in how COVID-19 was named: SARS-CoV 2, coronavirus) and may have adversely affected search performance for those who did not expand their queries to include such terms.

In fact, many of the successful runs from teams that used baseline runs from Anserini^30^ employed a query preprocessing tool produced by University of Delaware that used SciSpacy^31^ to lemmatize and remove non-stop words. Runs generated by Anserini comparing standard addition of various topic fields with and without the “Udel” method consistently showed improvements in retrieved relevant documents no matter what topics were used to construct the query and which indices were used.^32^ This approach was taken further either manually by certain teams (i.e., OHSU) or automatically, as seen in approaches in initial iterations of Covidex.^33^ In fact, adapting the queries to better represent document representations, or minimize query-document mismatch, has long been researched and includes work using relevance judgments^34^ and query expansion.^35^ Novel methods have focused on reverse: adjusting documentation representations to better represent queries - for example, Doc2Query^36^ was employed most commonly in Round 5 by one team, though this technique was not shown to be significantly associated with high performance in our study. However, the team that incorporated this technique submitted runs that were widely variable in performance, and may have used other features not found to be significant in our taxonomy. The importance of defining relevant terms in queries has also been reflected in human user studies, in which previous work by Hersh et. al has demonstrated the importance of search ability and domain knowledge of the user in a biomedical search task.^10^

### LIMITATIONS

This study had several limitations that future work could address. First, the instructions for describing methodologies in the run reports varied in detail. As such, the data used for this study were only as complete as what was provided in the reports. This not only presented a challenge to building our taxonomy, but also meant that important features may not have been (and likely were not) reported. In the future, teams should document methodologies that promote reproducibility or publish their results in reports as is done in the regular TREC challenges.

Second, it was difficult to capture run-specific differences between runs submitted by the same team, as team-specific features were often not provided. This had important implications in runs submitted in Round 5, where teams were allowed to submit up to 8 runs. While many runs submitted from the same team were largely similar (and often performed similarly), our methodology was not well-suited to capture nuances such as hyperparameter tuning that were likely small adjustments to otherwise similar methods and pipelines. We sought to characterize runs broadly, rather than capture each individual technique and adjustment in each run, since features built around individual techniques were subject to bias. However, to find a balance between granularity vs. breadth of techniques, we attempted to take into account differences between runs (even from the same team) using a one-hot encoded column of other techniques that we thought were unique enough to warrant specific inclusion. Future directions for this work may include identifying how to best capture adjustments between runs using similar techniques that result in different performances.

Third, our study was retrospective and limited in scope. While we studied techniques associated with performance on the CORD-19 dataset, these techniques may not be generalizable to other test collections. This was reflected in our work, where our regressions overfitted particularly performances with teams that used unique methodologies (i.e., associated a feature with significantly low or high performance despite a low number of teams employing this feature, such as ReQ-ReC^20^ or Proximity score). As mentioned above, implementation may have had a role in this “significantly” poor performance. Future work will be needed to prospectively evaluate these unique techniques across different developers and users.

## CONCLUSION

Using multivariate regression analysis, we developed and evaluated a taxonomy of features IR systems associated with high performance in the TREC-COVID Challenge. While our multivariate analysis demonstrates the utility of relevance feedback and the need for well-defined queries, it remains unclear which broad methodologies are associated with high performance in the TREC-COVID test collection. While our study has limitations in generating specific, prospective generalizations about IR systems and techniques, our work broadly showcases general techniques that may be useful in building search systems for COVID-19, and serves as a springboard for future work on TREC-COVID and related test collections.

## Data Availability

N/A

## Notes

### Competing Interest Statement

The authors have declared no competing interest.

### Funding Statement

We have no funding sources to disclose.

### Author Declarations

No IRB approval needed for this study of publicly available data from TREC-COVID.

